# Nanopore sequencing of influenza A and B in Oxfordshire and the United Kingdom, 2022-23

**DOI:** 10.1101/2023.11.21.23298840

**Authors:** Jennifer Cane, Nicholas Sanderson, Sophie Barnett, Ali Vaughan, Megan Pott, Natalia Kapel, Marcus Morgan, Gerald Jesuthasan, Reggie Samuel, Muhammad Ehsaan, Hugh Boothe, Eric Haduli, Ruth Studley, Emma Rourke, Ian Diamond, Tom Fowler, Conall Watson, Nicole Stoesser, Ann Sarah Walker, Teresa Street, David Eyre

## Abstract

**Objectives:** We evaluated Nanopore sequencing for influenza surveillance.

**Methods:** Influenza A and B PCR-positive samples from hospital patients in Oxfordshire, UK, and a UK-wide population survey from winter 2022-23 underwent Nanopore sequencing following targeted rt-PCR amplification.

**Results:** From 941 infections, successful sequencing was achieved in 292/388(75%) available Oxfordshire samples: 231(79%) A/H3N2, 53(18%) A/H1N1, and 8(3%) B/Victoria and in 53/113(47%) UK-wide samples. Sequencing was more successful at lower Ct values. Most same-sample replicate sequences had identical haemagglutinin segments (124/141;88%); a subset of samples also Illumina sequenced were very similar to Nanopore sequences.

Comparison of Oxfordshire and UK-wide sequences showed frequent inter-regional transmission. Infections were closely-related to 2022-23 vaccine strains. Only one sample had a neuraminidase inhibitor resistance mutation.

849/941(90%) Oxfordshire infections were community-acquired. 63/88(72%) potentially healthcare-associated cases shared a hospital ward with ≥1 known infectious case. 33 epidemiologically-plausible transmission links had sequencing data for both source and recipient: 8 were within ≤5 SNPs, of these, 5(63%) involved potential sources that were also hospital-acquired.

**Conclusions:** Nanopore influenza sequencing was reproducible and antiviral resistance rare. Inter-regional transmission was common; most infections were genomically similar. Hospital-acquired infections are likely an important source of nosocomial transmission and should be prioritised for infection prevention and control.

**Highlights:**

- Nanopore sequencing is a reproducible tool for influenza surveillance
- Inter-regional transmission of influenza was common across the UK
- Influenza anti-viral resistance was rare
- In 1 year most infections were genetically similar, hindering transmission studies
- Hospital-acquired infections are likely a key source of nosocomial transmission

## Introduction

Seasonal influenza infections cause significant mortality each year, as well as generating substantial peaks in healthcare demand.^1^ Vaccination is an important mitigation measure in those vulnerable to adverse outcomes arising from older age or comorbidities, as well as helping to control transmission and minimise rare but serious complications in children. Antiviral treatment and post-exposure prophylaxis with neuraminidase inhibitors are also used as part of a comprehensive approach to prevent severe influenza-related illness and death, alongside infection prevention and control measures.^2^

Whole genome sequencing of influenza allows surveillance of viral subtypes and potential resistance mutations. It also potentially enables early warning of divergence of circulating strains from those selected for seasonal vaccines, which in turn can be used to help plan control interventions, as well as future vaccine design. Sequencing could also help direct treatment guideline recommendations, or even individual treatment, in the context of antiviral resistance. Sequencing also has potential as an infection prevention and control tool, helping to identify, and hence interrupt, healthcare-associated influenza transmission. Furthermore, sequencing can also allow detection of pathogens that have undergone genetic changes, which may reduce sensitivity of other molecular assays.^3^

Primers targeting conserved sequences across all eight segments of the influenza A genome have been used to facilitate sequencing using a variety of techniques.^4^ Most recently, approaches using targeted^5,6^ Nanopore sequencing have been described, including comparisons of accuracy with Illumina sequencing,^6^ although Nanopore sequencing technology has developed further since. Successful reconstruction of influenza genomes from Nanopore metagenomic^7,8^ sequencing is possible, albeit mostly with higher viral loads. Sequencing has been used to detect circulating, subtypes, antiviral resistance mutations, and hospital-based clusters.^8–10^

Here we combine analysis of whole genome sequencing of influenza from a large UK teaching hospital group, providing services to around 1% of the UK population, with sequences from a representative UK-wide household study, from the winter 2022/2023 season. We describe an approach for applying Nanopore technologies including a rigorous quality control pipeline to ensure high quality sequencing. We identify evidence of frequent inter-regional transmission in the UK, as well as an important role for hospital-acquired influenza infections in onward transmission in healthcare.

## Materials and methods

### Samples, settings and ethical approvals

Residual nasal and/or throat swab samples that had tested positive for influenza A or B by PCR-based GeneXpert assay (Cepheid) or Biofire FilmArray (bioMérieux) were collected from the microbiology laboratory of Oxford University Hospitals (OUH). OUH consists of 4 teaching hospitals with a total of ∼1100 beds, providing secondary care services to a population of 750,000 in Oxfordshire, UK, and specialist services to the surrounding region. Cycle threshold (Ct) values, indicative of viral loads in swab media, were available for the GeneXpert assay; the Biofire platform provides only a binary result. Ethical approval for use of the samples, and linked de-identified hospital record data, was granted by the London-Queen Square Research Ethics Committee (17/LO/1420).

Influenza A/B positive samples from a pilot extension to the Office for National Statistics COVID-19 Infection Survey (CIS) were also obtained. CIS was a large longitudinal household survey, broadly representative of the wider UK population, conducting PCR tests for SARS-CoV-2 on self-collected nose and throat swabs and collecting questionnaire data approximately monthly. The study received ethical approval from the South Central Berkshire B Research Ethics Committee (20/SC/0195). From October 2022, a random subset of ∼750 swabs received per week were additionally tested by multiplex PCR (ThermoFisher TaqPath™ COVID-19, Flu A/B, RSV ComboKit A49867); PCR results, symptoms and temporal patterns have been described previously.^11^ Samples positive for influenza A/B were sent to the University of Oxford for sequencing, together with PCR results and sample metadata (age provided as groups).

### Nucleic acid preparation, RT-PCR, and sequencing

Nucleic acid was extracted for all samples using the Kingfisher Flex (ThermoFisher) MagMax TaqPath protocol using 200µl sample and eluted in 50µl elution buffer. For the CIS samples, influenza A and B were differentiated using the RealStar Influenza RT-PCR Kit (Altona Diagnostics). Purified RNA underwent targeted RT-PCR for influenza A or B. Sequencing of resulting cDNA was performed, multiplexing up to 40 samples, using the Nanopore GridION device. A subset of sequences also underwent Illumina sequencing to evaluate the accuracy of Nanopore sequencing (see Supplement for details).

### Bioinformatic and statistical analysis

The Nanopore bioinformatics workflow was written in nextflow^12^ and packaged in conda; it is publicly available on gitlab here https://gitlab.com/ModernisingMedicalMicrobiology/flu-workflow. Competitive mapping of sequence reads was used to identify influenza subtypes and to generate consensus genomes for analysis. The influenza model in IRMA (https://wonder.cdc.gov/amd/flu/irma/) was used to assemble raw Illumina reads into consensus sequences to assess Nanopore sequencing accuracy (see Supplement).

To address potential misidentification of samples arising from low level laboratory contamination, barcode cross-over effects arising from imperfect demultiplexing of reads, and Nanopore sequencing errors, we developed several filters described in the Supplement (Figures S1, S2).

Phylogenetic trees were created based on alignments of all successfully sequenced haemagglutinin (HA) segments for A/H3N2, A/H1N1, and B/Victoria sequences. Strains for 2022-23 northern hemisphere vaccines^13^ were included in the trees, with one randomly selected as an outgroup to root the tree. Similarly, phylogenetic trees were created based on neuraminidase (NA) segments, and for whole genomes. Pairwise SNPs between segment and whole genome sequences were obtained from the cophenetic distances extracted from trees multiplied by the alignment length. Rates of influenza HA segment mutation were estimated from sample dates and sequence alignments using BEAST (version 1.10.4) (see Supplement).

Multi-variable logistic regression was used to investigate associations with sequencing success, using sequencing of the HA segment as the outcome, and Ct values, input cDNA concentration and sub-study (Oxfordshire vs. UK-wide) as exposures.

We used a permutation test to assess for evidence of regional clustering of influenza genomes. The median SNP difference between sequences from Oxfordshire was compared to the median difference between sequences where one was from Oxfordshire and the other from the UK-wide sequencing dataset. To assess if the observed differences were compatible with chance we permuted the sequence location labels 1000 times, and compared the distribution of the observed SNP differences to this random distribution.

We searched each NA segment for previously reported mutations conferring resistance to neuraminidase inhibitors, i.e., the H275Y mutation in A/H1N1pdm09 viruses linked to oseltamivir resistance, and the I223R mutation conferring both oseltamivir and zanamivir resistance. We also searched for E119V and R292K in A/H3N2.

### Analysis of healthcare-associated infections

Influenza infections diagnosed within ≤72 hours of admission were deemed community-associated, and those diagnosed after >72 hours to be potentially-healthcare associated.^14^

We searched for evidence of healthcare influenza transmission by considering cases potentially infectious from 2 days before a positive test to 5 days afterwards,^15^ and cases at risk of acquisition from 7 to 2 days^16^ before their positive test, allowing for a short delay between symptom onset and testing. Under these illustrative assumptions set to prioritise sensitivity for detecting transmission events (i.e., using upper limits of infectious/incubation periods), possible transmission events were defined where potentially healthcare-associated cases were present on the same hospital ward while at risk as an infectious case of the same influenza type (as defined by the initial diagnostic PCR, i.e. A or B). The plausibility of these potential transmission links was also evaluated by sequencing where both the case and contact’s infections were successfully sequenced. Infections within HA segments within ≤5 SNPs were considered to represent potential transmission (see Results).

## Results

### Oxfordshire samples and sequences

Between 01 August 2022 and 31 March 2023, 996 influenza A or B positive samples were identified by the OUH microbiology laboratory representing 941 infections in 940 unique patients; 931 influenza A (93%), 61 influenza B (6%), and 4 both influenza A and B (<1%, all in children of ages eligible to have received live attenuated influenza vaccine, LAIV). The median (IQR) patient age across all positive results was 50 (24-76) years (range <1 to >100 years), 549 (55%) were from female patients.

508 samples were retrieved by the research laboratory, 388 of which had sufficient residual sample available for sequencing (370 influenza A, 17 influenza B, 1 mixed) (Figure S3A). Ct values were available for 308 samples, median 24.2 (IQR: 20.3-27.6). Successful sequencing of at least 2 segments enabled identification of the influenza type in 292/388 (75%) samples: 231 (79%) A/H3N2, 53 (18%) A/H1N1pdm09 (hereafter A/H1N1), and 8 (3%) B/Victoria. The HA segment was successfully sequenced in 269/388 (69%) of available samples, and these sequences were used in phylogenetic comparisons.

### UK-wide samples and sequences

Within the CIS respiratory pilot, 14,939 randomly selected swabs from 14,664 unique participants were tested between 10 October 2022 and 26 February 2023. In total 130 samples were positive for influenza A/B (positivity=0.9%); these positive samples were obtained from study participants in England (n=106), Scotland (n=11), Northern Ireland (n=8), and Wales (n=5). Most participants had been vaccinated in the current and/or previous influenza season (n=90, 69%), most reported some form of symptoms at the point the swab was taken (n=86/124, 69%, data missing for 6, see ^11^ for details), there was an equal split of participant sexes (n=65 female/male) and a wide spread of ages ranging from ≤5 to ≥80 years, median range 30-40 years (IQR: 10-20 years, 50-60 years) (only age groups available). Median (IQR) Ct values on initial testing (combined influenza A/B) were 29.9 (27.3-32.1).

Following transport of samples to the sequencing laboratory, on repeat PCR testing 87/130 samples yielded a subtype (5 other samples had initial Ct >30, did not yield a subtype and sequencing was not attempted; the remaining 38 were PCR negative on re-testing with a different assay) (Figure S3B): 80 (92%) were influenza A, 5 (6%) influenza B, and 2 (2%) both (1 in a child vaccinated on the same day, presumed to have received LAIV, and the other in an unvaccinated adult). Despite some samples testing PCR-negative on repeat testing, the Ct values were otherwise like the original results, median (IQR) 29.6 (26.7-31.1).

113 samples had sufficient material for sequencing. This included 33 samples that tested PCR negative on repeat testing, but where sequencing was attempted anyway in case small quantities of degraded RNA was still present (using both influenza A and B primers to 31 December 2022, and only influenza A primers thereafter). Overall, 53/113 (47%) had a flu subtype identified from sequencing: 41 (77%) A/H3N2, 11 (21%) A/H1N1, 1 (2%) B/Victoria. Successful HA segment sequencing was achieved in 50/113 (44%). Only 1/33 (3%) samples negative on repeat PCR testing produced a HA segment sequence, hence in samples PCR-positive on repeat testing the success rate was higher, 49/80 (62%).

### Negative, control and replicate samples

A further 22 influenza A and B negative patient samples from Oxfordshire were sequenced, as well as 58 water controls, none were positive by sequencing, i.e., none had any segments that passed quality control filters.

As part of internal quality assurance, some samples were sequenced more than once. Overall, 705 sequencing attempts were made for 501 samples across both datasets; 433 (61%) yielded a sequence including an HA segment that passed quality filters. The same sample was successfully sequenced twice (n=63), three times (n=16), or four times (n=5), providing a total of 141 replicate pairs for comparison.

### Characteristics of successfully sequenced samples

Influenza qPCR Ct values were available for 388 samples where sequencing was attempted (541 sequencing attempts). The percentage of samples with a successfully sequenced HA segment was highest with low Ct values, i.e., high viral loads, e.g., >80% for Ct values ≤25, 67% for >25-≤30, and 28% for Ct values >30-≤35; only one of 15 (4%) attempts with Ct >35 yielded a successful sequence (Figure 1A). Similarly, samples with high total cDNA concentrations after reverse transcription were more likely to be successfully sequenced, e.g., 27% with ≤25 ng/µl compared to 95% with >100 ng/µl (Figure 1B).

**Figure 1.**
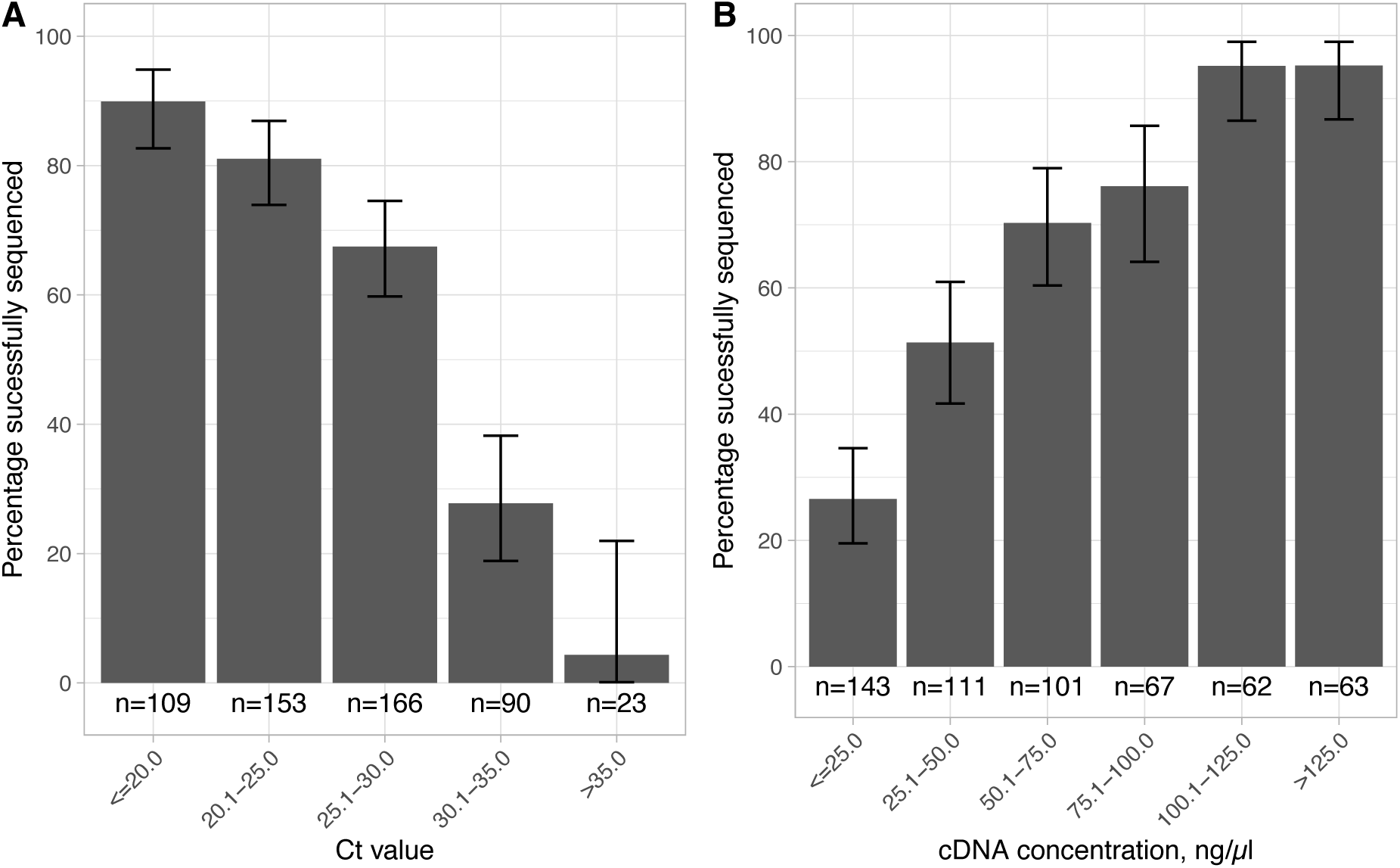
Relationship between sequencing success and Ct values and cDNA concentrations. 541 attempted sequences had an associated Ct value and 547 an associated cDNA concentration recorded (sequencing was attempted more than once for some samples). Error bars show 95% exact binomial confidence intervals.

Using multivariable logistic regression, both lower Ct values (adjusted odds ratio, aOR=0.84 per unit higher [95%CI 0.79-0.89; p<0.0001]) and higher cDNA concentrations (aOR=1.30 per 10ng/µl higher [1.20-1.41; p<0.0001]) were independently associated with sequencing success. Oxfordshire samples were less likely to be successfully sequenced at a given Ct value and cDNA concentration (aOR=0.44 [0.23-0.86]), likely reflecting the fact that different PCR assays and hence different Ct value scales were used in each sampling group. Taken together, the most likely explanation for the higher proportion of samples with a successfully sequenced HA segment in the Oxfordshire dataset (69% vs 62% in the samples PCR-positive on repeat testing in UK-wide study) was the lower Ct values, i.e. higher viral loads, in these symptomatic patients presenting to hospital, compared to the community samples obtained from UK-wide participants as part of a systematic sampling survey (Figure S4).

Across 141 pairs of replicate sequences and considering HA segment differences, most pairs were identical (n=124, 88%); the remainder were 1 SNP different (n=13, 9%), 2 SNPs different (n=2, 1%), or 4 SNPs different (n=2, 1%) (Figure 2A). In 124 pairs with ≥60% of the whole genome identified in both samples, most SNP differences were ≤10 (115, 93%; Figure 2B). For 6/9 of these pairs with >10 SNP differences, these differences were ≤10 SNPs in a sensitivity analysis excluding segments S2 and S3 which had a greater tendency for cross mapping between influenza types (Figure S5). Comparing HA segments between successfully sequenced pairs of different samples from the same influenza subtype from across Oxfordshire and England in the same winter season, up to ∼50 SNPs were seen (Figure 2C). Comparing genomes based on all eight segments up to ∼800 genome-wide SNPs were seen (Figure 2D).

**Figure 2.**
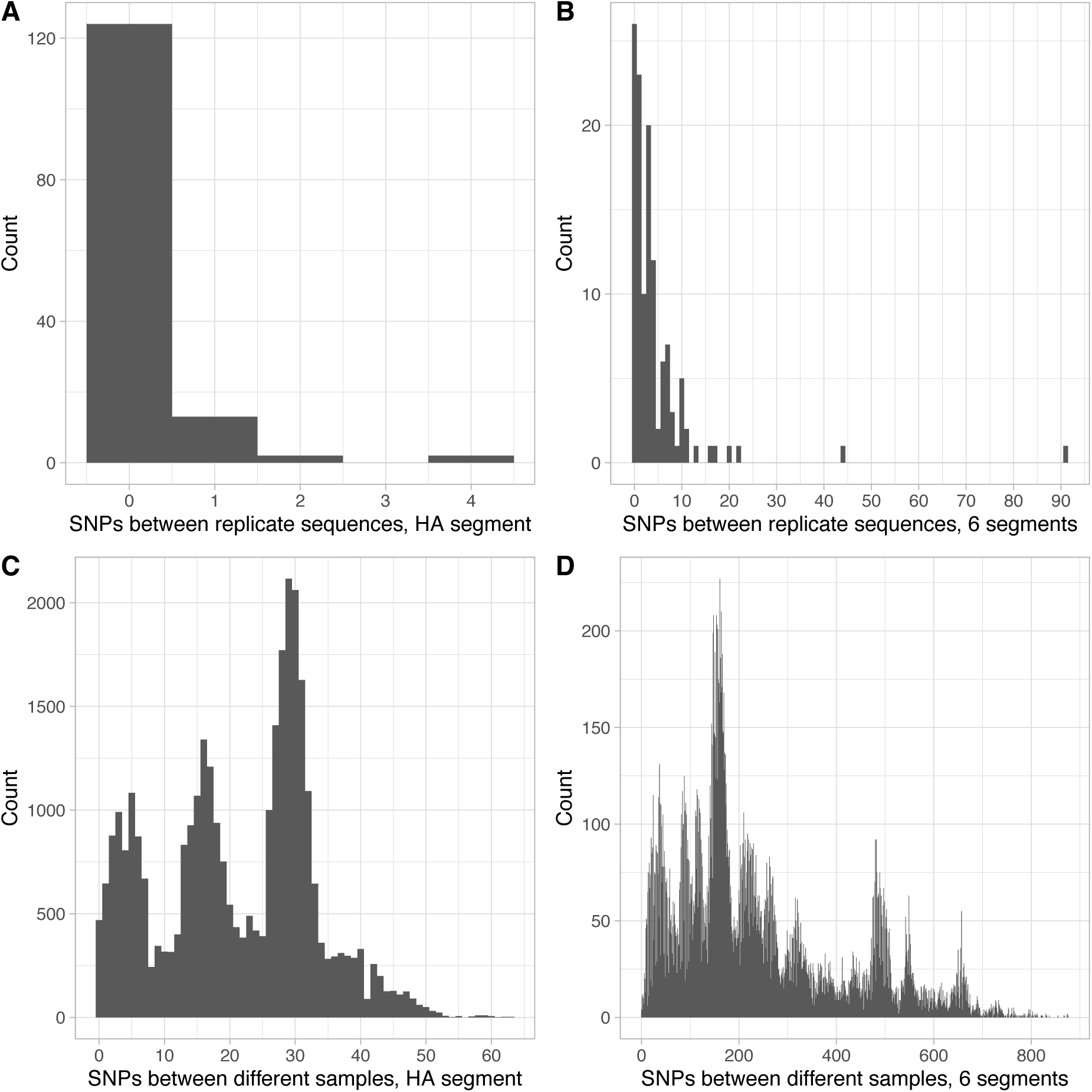
Distribution of single nucleotide polymorphism (SNPs) between replicate and different sample pairs. Panels A (n=141) and C (n=32,992) show differences for the HA segment and B (n=124) and D (n=29,147) for whole genomes based on all 8 segments (see Figure S5 for sensitivity analysis using only six segments that removes several high SNP replicates; segments S2 and S3 had a greater tendency to cross mapping between flu types).

Although the primers used allow for amplification of all 8 influenza segments, coverage of each segment varied, with segments 4 (HA), 7 (M1 M2), and 8 (NEP NS1) best covered in influenza A (Figure 3).

**Figure 3.**
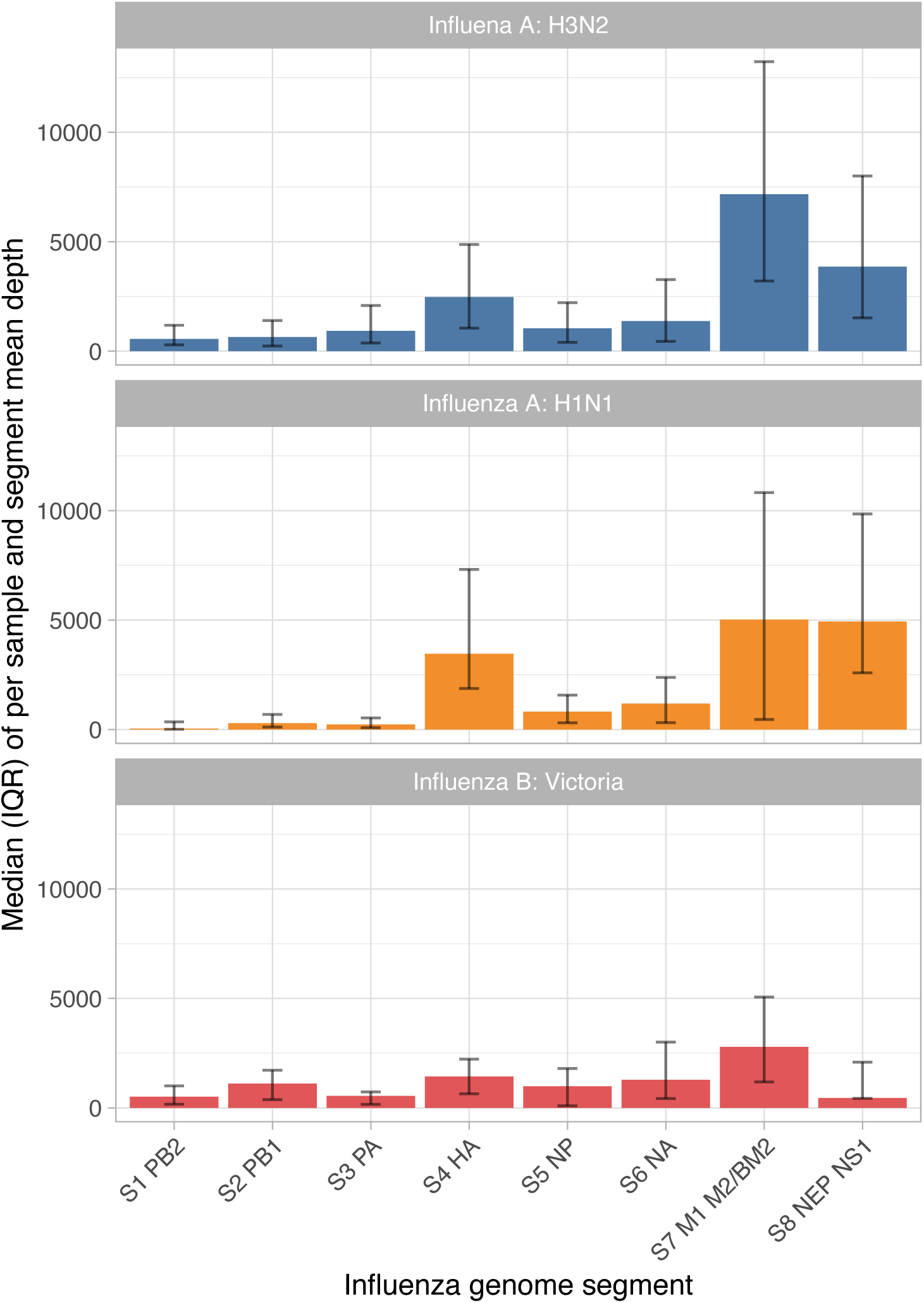
Depth of coverage by influenza genome segment. Mean depth was calculated for each segment for each sample in which the HA segment was successfully sequenced, i.e., the sequences used in phylogenetic analyses. The median of the mean depth values for each influenza type and segment is shown. Error bars show interquartile ranges, IQRs.

### Comparison of Illumina and Nanopore sequencing results

A total of 26 samples had HA segments that were successfully sequenced by both Illumina and Nanopore methods. As some samples were successfully Nanopore sequenced more than once, there were 51 Illumina vs Nanopore comparisons possible: 30 (59%) pairs were 0 SNPs different, 7 (14%) were 1 SNP different, and 14 (27%) were 2 SNPs different. In 23 samples with ≥2 Nanopore sequences the number of SNPs between the replicates and the matching Illumina sequence was the same for 22 (96%), i.e., any differences between Illumina and Nanopore were reproduced across multiple Nanopore replicates, potentially consistent with systematic Nanopore sequencing errors.

### Phylogenetic analysis of Oxfordshire and UK sequences

HA sequences from UK-wide A/H3N2 and A/H1N1 samples were distributed throughout phylogenetic trees of Oxfordshire samples (Figure 4; similar findings from alignments of six segments excluding S2 and S3, Figure S6), indicative of relatively frequent inter-regional transmission. There was some evidence for limited local clustering, e.g., within A/H3N2 HA sequences the median pairwise SNP differences between Oxfordshire sequences was 22.3 (IQR 11.8-29.8), compared to 26.6 (13.8-30.9) in Oxfordshire-UK pairs and 27.7 (14.9-33.0) in UK-UK pairs (permutation test p value for the observed ratio of Oxfordshire-Oxfordshire / Oxfordshire-UK SNP differences being at least as great as observed by chance = 0.015).

**Figure 4.**
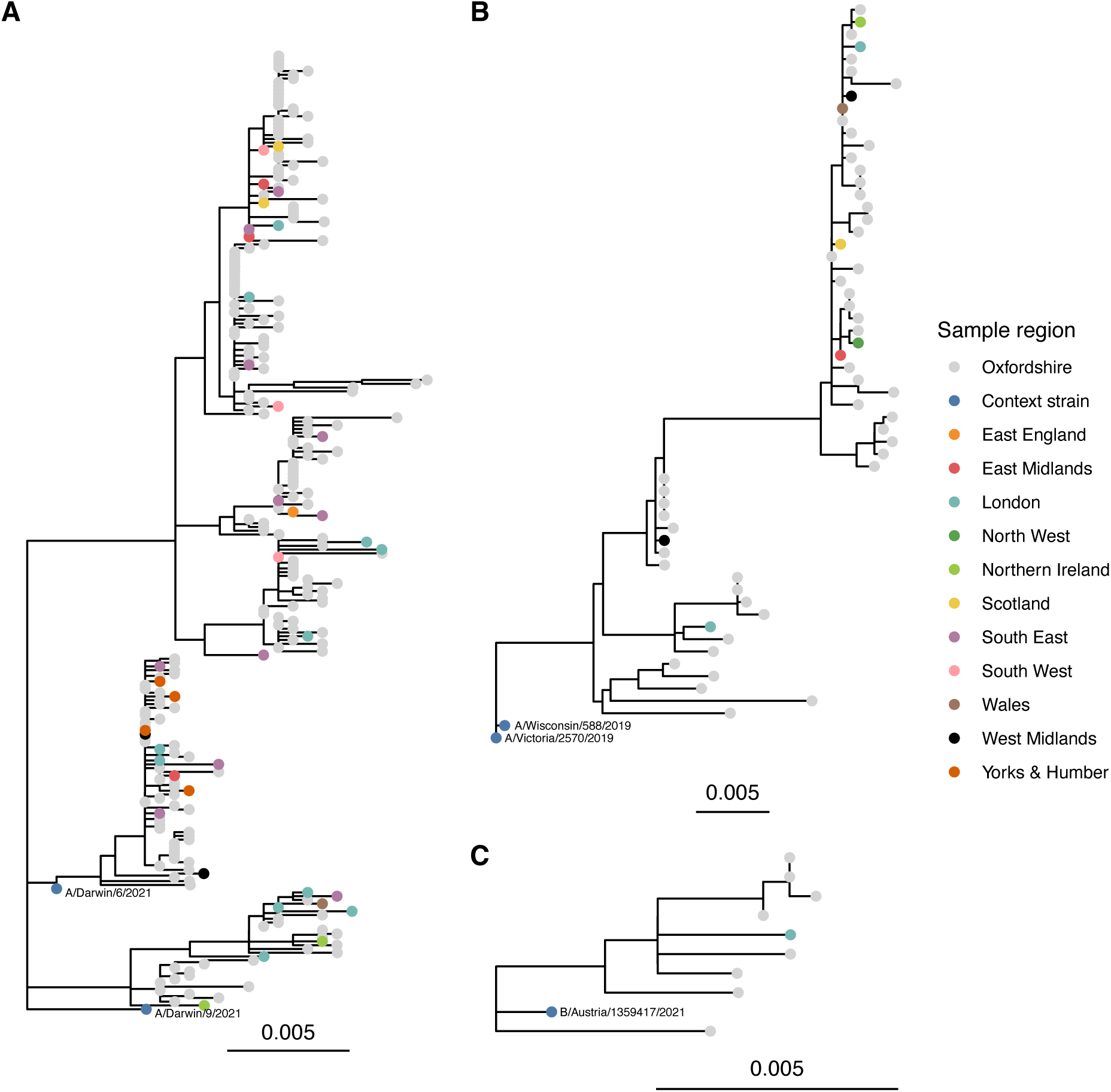
Maximum likelihood phylogeny of HA segment sequences for influenza A H3N2 (panel A), H1N1 (panel B), and influenza B/Victoria (panel C). Sequences are coloured according to source dataset, 2022-23 northern hemisphere vaccine strains are shown for context, with one strain used as an outgroup to root the tree. (See Figure S6 for sensitivity analysis excluding segments S2 and S3 which had a greater tendency for cross mapping between influenza types.)

Sequences were relatively closely related to the 2022-23 northern hemisphere WHO recommended vaccine strains,^13^ with a median (IQR) nucleotide diversity over the length of the HA segment between vaccine strains and sequences from the combined Oxfordshire/UK-wide datasets of 0.012 (0.010-0.015), 0.024 (0.016-0.025), and 0.007 (0.006-0.007) for A/H3N2, A/H1N1, and B/Victoria respectively. Results were similar for the NA segment, 0.010 (0.007-0.017), 0.012 (0.011-0.013), and 0.006 (0.005-0.008) respectively.

### Drug resistance and evolutionary rates

Only one sample had evidence of a mutation associated with resistance to neuraminidase inhibitors. A sample from a 59 year old female patient from Oxfordshire in January 2023 had a H275Y mutation in an A/H1N1 infection. We estimated the rate of substitutions in the HA segment of A/H3N2 to be 6.3 x 10^-3^ per base per year (95% credibility interval 4.6-8.0 x 10^-3^), and 6.3 x 10^-3^ (3.6-9.2 4.6-8.0 x 10^-3^) per base per year for A/H1N1. This corresponds to approximately 11 SNPs per segment per year.

### Oxfordshire cases vs ward contacts

The 996 influenza A or B positive samples identified by the OUH microbiology laboratory represented 941 infections in 940 unique patients (one patient had an influenza B infection 53 days after testing influenza A positive). Although serial samples were obtained from 42 patients (median [IQR] 8 [1-12] days between first and last samples), these were not routinely sequenced; only 2 pairs (obtained ≤1 days apart) were sequenced with 0 and 4 SNPs between HA segments and 10 and 8 SNPs between six segment alignments (excluding S2 and S3, as above).

Considering the 941 infections, 849 (90%) were community acquired (diagnosed within 72 hours of admission), 4 (0.4%) were compatible with LAIV, and the remaining 88 (9%) were potentially healthcare-associated,^14^ 33 (4%) diagnosed within 3-7 days of admission, and 55 (6%) after >7 days of admission.

Under assumptions set to prioritise sensitivity to detect most potential transmission, 63/88 (72%) of potentially healthcare-associated cases had a link with ≥1 known infectious case on the same hospital ward while at risk. There were multiple possible sources identified for some infections, such that there were 359 potential transmission links overall. Some cases were exposed to multiple patients in admission or ambulatory units, others formed ward-based clusters, while other potential transmission events involved only a pair of cases (Figure 5).

**Figure 5.**
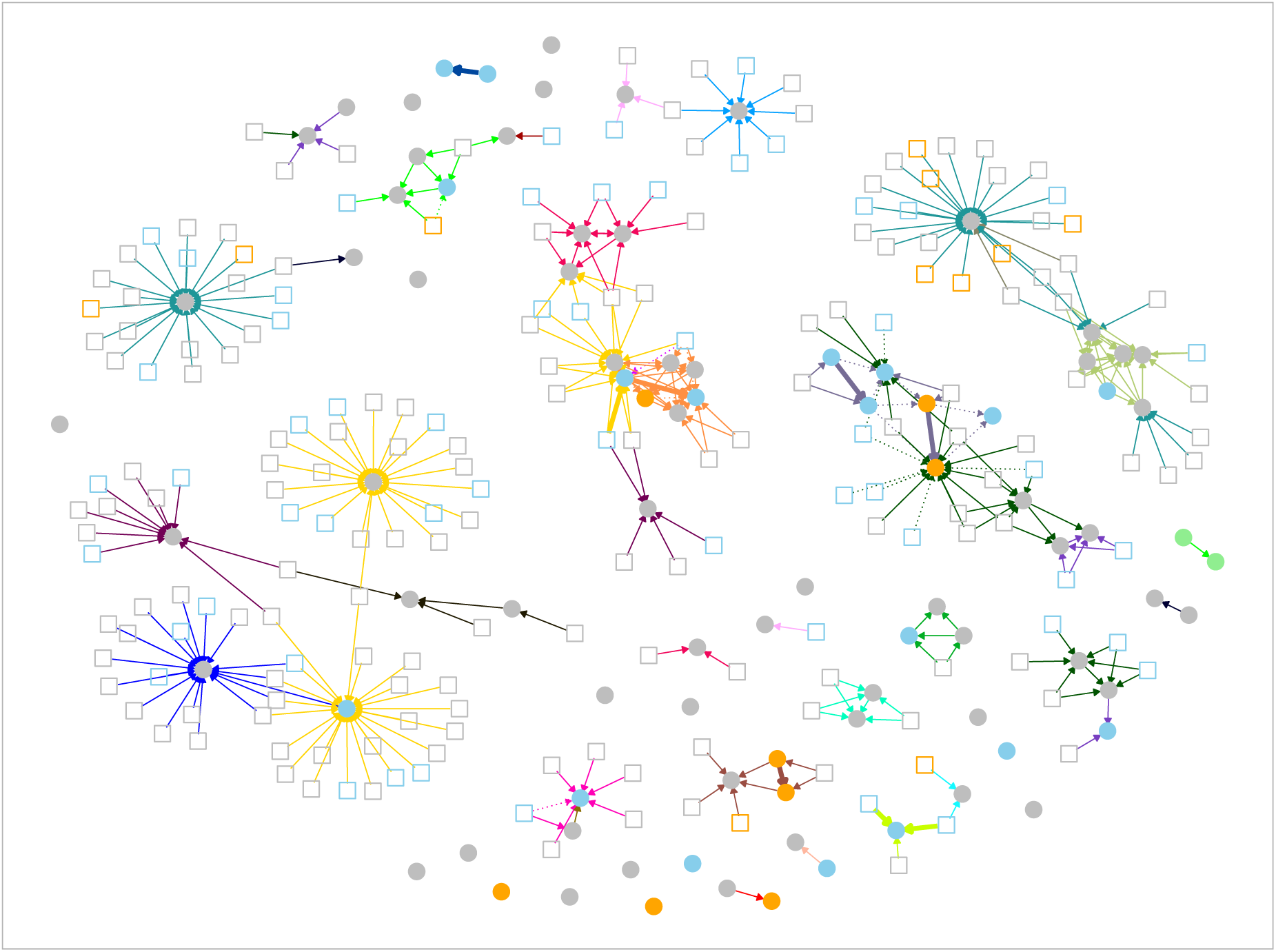
Plausible sources for 88 potential healthcare-associated influenza infections throughout the study period, based on ward overlap during defined infectious and incubation periods. Infections acquired after >72 hours as a hospital inpatient are shown as filled circles, community-associated cases are shown as squares. Nodes are coloured by influenza subtype as determined from sequencing: A/H3N2 in blue, A/H1N1 in orange, B/Victoria in green, unknown (not sequenced or not sequenced successfully) in grey. Possible transmission events, shown as edges, are drawn where potentially healthcare-associated cases were present while at risk on the same hospital ward as an infectious case of the same influenza type (as defined by the initial diagnostic PCR, i.e. A or B). The edges are coloured according to the ward on which contact occurred. The yellow edges and blue edges represent acute medical admissions units at two hospitals and the teal edges those occurring on an acute medical ambulatory assessment unit. Orange, red, pale green, and lime green clusters occurred on acute medical / geratology wards. The turquoise cluster represents a paediatric ward, the pink cluster maternity wards, and the brown cluster a haematology ward. Thick solid edges are where transmission is supported by sequencing, i.e., ≤5 SNPs between cases, dotted lines indicate where sequencing makes transmission less likely either due to >5 SNPs (all actually ≥15 SNPs) or different influenza subtypes.

Of 359 potential transmission links, 33 had sequencing data available for both the putative transmission donor and recipient: 13 (39%) links had different flu subtypes excluding transmission, 4 had matching subtypes but the HA segment was not identified in at least one infection, five were 0 SNPs different, one was 1 SNP, one 2 SNPs, one 5 SNPs, and the remaining eight were ≥15 SNPs different (Figure 5). Given the error rate in replicates and estimated of HA segment evolution of 11 nucleotides/year in A/H3N2, if up to 5 SNPs were considered consistent with recent transmission in the same influenza season, 8/16 (50%) assessable transmission pairs with the same flu subtype were within this threshold, and 7/16 (44%) within ≤2 SNPs. This compares to the 3593/24376 (15%) of all pairs of Oxfordshire sequences of the same flu type within ≤5 SNPs and 1520 (6%) within ≤2 SNPs.

Amongst the 8 cases with ≤5 SNPs between them, 5 (63%) involved potential donors that were also hospital acquired (all within ≤2 SNPs), highlighting that these cases may be an important source of infection, despite accounting for only 88/931 (9%) cases overall.

Most Oxfordshire infections occurred in December 2022, accompanied by a rise in the proportion of cases that were healthcare-associated from the second week of December onwards (Figure 6A/6B). Clustering genomically similar infections (i.e., those within ≤5 SNPs of another case), most community and healthcare associated cases formed part of a small number of large clusters (3 for A/H3N2 and 1 for A/H1N1; Figure 6C). The large number of similar community and healthcare associated infections means identifying hospital transmission using genomics alone is unlikely to feasible, necessitating combined analysis with ward contact data as above.

**Figure 6.**
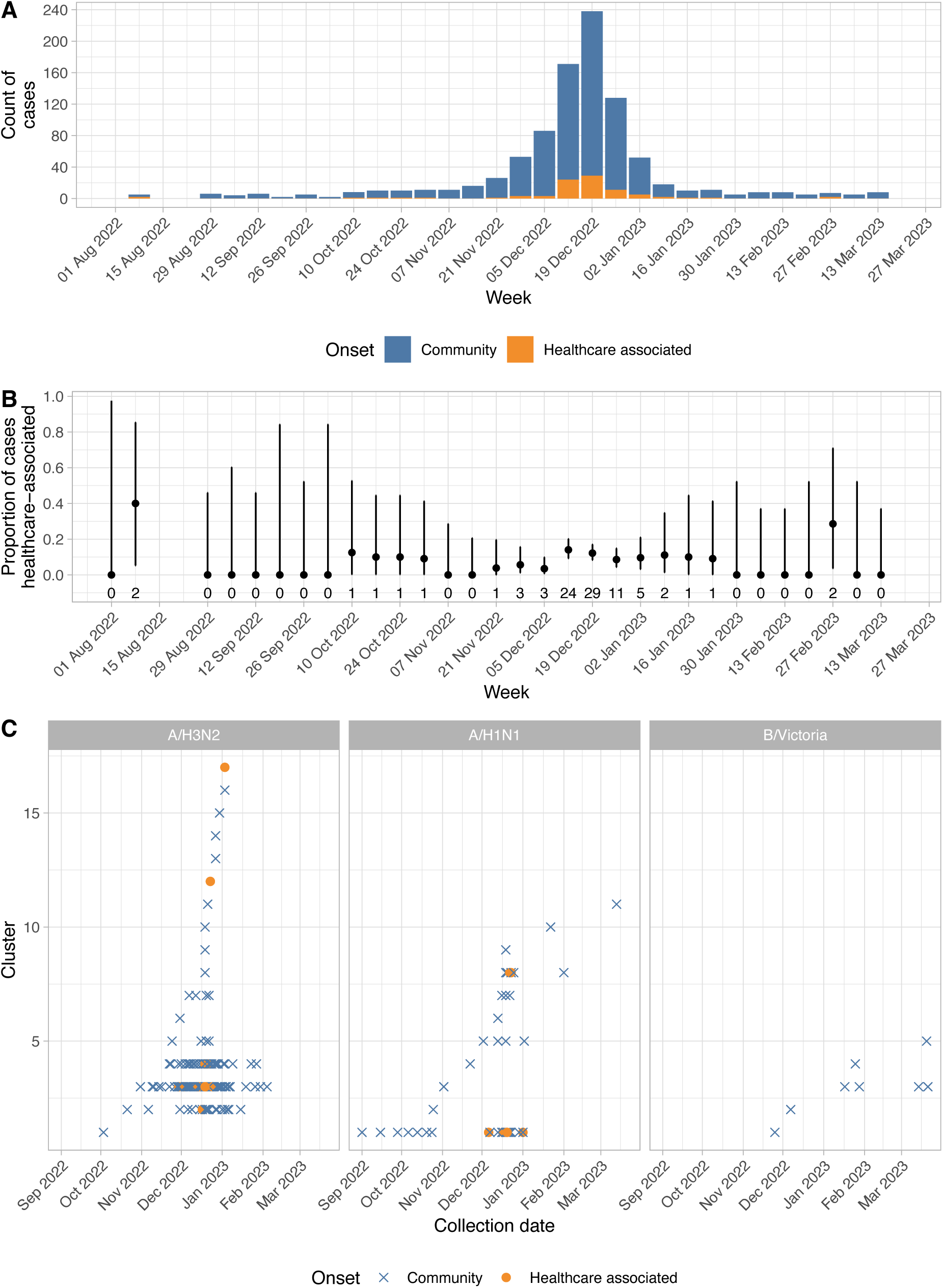
Epidemiology of Oxfordshire influenza. Panel A shows hospital diagnosed infections (n=941), classified by community onset (diagnosis ≤72 hours of hospital admission) vs. potentially healthcare-associated (>72 hours). Panel B shows the proportion of cases that were potentially healthcare-associated, error bars indicate 95% binomial confidence intervals, and the numbers beneath each point the number of potentially healthcare-acquired cases per week. Panel C shows successfully sequenced infections with an HA segment called (n=269), each case >5 HA segment SNPs different all previous cases is shown on a new horizontal line.

## Discussion

We show it is possible to undertake Nanopore-based sequencing of influenza at scale throughout a winter influenza season. While we show good concordance between circulating and vaccine strains for A/H3N2, A/H1N1, and B/Victoria and a low frequency of anti-viral resistance, our approach could also be used to provide surveillance in future seasons where this might not be the case.

We show, as has been identified previously for influenza and other respiratory viruses,^7,17^ that sequencing success depends on the amount of viral RNA present. Ct values >30 were associated with a reduced chance of sequencing success, and our efforts to sequence samples that had very high Ct values or that had potentially degraded in transport show there is little point in attempting sequencing where repeat PCR is negative or Ct values are >35. Given the distribution of Ct values observed in our hospital group, this means that attempting to comprehensively sequence all influenza in a hospital is not possible. In our study this was compounded by over half of samples not being retrieved for sequencing due to logistical constraints, or not having sufficient residual sample for sequencing.

Nevertheless, we are still able to make several observations. Firstly, there was only limited geographic structure when comparing sequences from Oxfordshire infections to those from across the UK. This is consistent with frequent transmission between Oxfordshire and multiple parts of the UK, i.e. widespread dissemination across the UK. However, as we did not have a larger UK-wide sample, we cannot exclude geographic structure elsewhere, e.g., it is possible that other specific locations elsewhere in the UK may experience less transmission with the rest of the UK than is seen in Oxfordshire.

Secondly, many community and hospital-associated infections occurring concurrently were genomically very similar. This limited genomic diversity across a few key weeks of a winter season means that genomic data need to be combined with hospital ward movement data if inferences about hospital-based transmission are to be attempted; sequencing data alone is insufficient given the limited background diversity. We were, however, able to refute some apparent potential transmission of influenza in hospitals using sequencing. We also show that genomically and epidemiological plausible sources of transmission are enriched for cases that are themselves likely hospital-acquired. This same pattern has also been observed for SARS-CoV-2 where patients are most infectious shortly after they are initially infected, such that nosocomial cases account for much hospital-based transmission.^18,19^ Focusing infection control efforts around these cases is an important priority. It should also be considered that genomically and epidemiologically linked pairs, could also represent ‘co-secondary’ cases, where both are acquired from a common, but unascertained source.

We show that Nanopore sequencing can produce accurate results compared to Illumina technology, and that results were largely reproducible across replicate sequences. Inclusion of repeated samples on multiple sequencing runs provided important internal quality assurance. We show that it is possible to infer influenza subtypes using other segments, even where the HA and NA segments are not both sequenced. We attempted to sequence samples even with very low viral loads, and although we normalised input cDNA concentrations as much as possible when multiplexing samples, inevitably samples with low concentrations accounted for a lower proportion of the total cDNA input. This meant that such samples were disproportionally at risk of contamination – both in the laboratory and from errors demultiplexing reads (which is an imperfect process). We developed filters that were able to largely mitigate this; some required absolute levels of data to be generated to call a specific segment as present, while others required a proportion of the data from the whole run or sample to be associated with a specific segment subtype in the sample. It is probable that if stricter requirements were made excluding low viral load samples from sequencing attempts, enabling tighter normalisation of cDNA concentrations, that these filters may be less critical. While the primers we used amplified all 8 segments, the efficiency across segments varied, as has been reported previously.^6^ For samples with sufficient input virus this can be overcome by a large enough overall sequencing depth, but this did limit whole-genome reconstruction in samples with lower viral loads.

Other studies utilising both Oxford Nanopore Technologies and Illumina platforms have investigated influenza transmission,^10,20,21^ focusing on specific hospital wards or known outbreaks. Including influenza infection cases from across a hospital setting in this study allowed the relative contribution of community vs hospital-onset cases to transmission to be assessed. Additional nationwide sequences allowed the relative extent of regional vs. national transmission to be studied. Going forward this will be useful to determine whether a local snapshot of circulating virus is representative of infections in the wider population.

Our study has several limitations. Some of the most important are covered above, namely that we only attempted sequencing on a subset of positive samples and that sequencing success rates reduced at lower viral loads. Whilst this meant that only a relatively small subset of potential transmission links could be investigated using sequencing in the Oxfordshire hospital samples, this is likely to apply to all studies attempting to use sequencing to recover transmission links. Several UK-wide study samples degraded in transport reducing the number available for sequencing. Coverage of whole genomes was only partial, so despite a whole-genome approach, much of our analysis is focused on the HA segment. The PCR assays used for the Oxfordshire and UK-wide studies differed (as did sample handling conditions) meaning that Ct values are not directly comparable across studies. Whilst sequencing of samples was undertaken in general within a week of collection for Oxfordshire samples, analysis was completed when the whole dataset was collected meaning influenza surveillance was not timely for the purposes of transmission and outbreak detection. Future work should attempt to improve yields from lower viral load samples and consider if primer designs can be modified to produce more uniform coverage across the whole genome. It would also be of interest to compare community and hospital samples from the same location, to look for any differences, and this may also be relevant when considering surveillance programme designs.

In conclusion, we have developed a method for high-throughput Nanopore sequencing of influenza A and B viruses. This has potential to be used as a diagnostic where subtyping or resistance detection is important, for surveillance, and as an infection control tool. Healthcare-associated influenza cases are likely to be an important source of onward transmission in hospitals and infection prevention and control efforts around these patients should be prioritised.

## Data availability

All genomes generated during this study are publicly available under European Nucleotide Archive Project PRJEB56915, https://www.ebi.ac.uk/ena/browser/view/PRJEB56915.

Oxfordshire patient data analysed are available from the Infections in Oxfordshire Research Database (https://oxfordbrc.nihr.ac.uk/research-themes-overview/antimicrobial-resistance-and-modernising-microbiology/infections-in-oxfordshire-research-database-iord/), subject to an application and research proposal meeting on the ethical and governance requirements of the Database.

De-identified COVID-19 Infection Survey study data are available for access by accredited researchers in the ONS Secure Research Service (SRS) for accredited research purposes under part 5, chapter 5 of the Digital Economy Act 2017. For further information about accreditation, contact or visit the SRS website.

## Transparency declaration

No author has a conflict of interest to declare.

## Acknowledgements

We wish to thank all the individuals who participated in the COVID-19 Infection Survey. We are grateful for the support of all the COVID-19 Infection Survey team:

Office for National Statistics: Sir Ian Diamond, Emma Rourke, Ruth Studley, Nick Taylor, Tina Thomas, Fiona Dawe; Office for National Statistics COVID Infection Survey Analysis and Operations teams in particular Dawid Pienaar, Joy Preece, Sarah Crofts, Lina Lloyd, Michelle Bowen, Daniel Ayoubkhani, Russell Black, Antonio Felton, Megan Crees, Joel Jones, Esther Sutherland;

University of Oxford, Nuffield Department of Medicine: Ann Sarah Walker, Derrick Crook, Philippa C Matthews, Tim Peto, Emma Pritchard, Nicole Stoesser, Karina-Doris Vihta, Jia Wei, Alison Howarth, Kevin K Chau, Lucas Martins Ferreira, Brian D Marsden, Wanwisa Dejnirattisai, Juthathip Mongkolsapaya, Sarah Hoosdally, Richard Cornall, David I Stuart, Gavin Screaton;

University of Oxford, Nuffield Department of Population Health: Koen Pouwels;

University of Oxford, Big Data Institute: David W Eyre, Katrina Lythgoe, David Bonsall, Tanya Golubchik, Helen Fryer;

University of Oxford, Radcliffe Department of Medicine: John Bell;

Oxford University Hospitals NHS Foundation Trust: Stuart Cox, Kevin Paddon, Tim James; University of Manchester: Thomas House;

UK Health Security Agency: Julie Robotham, Paul Birrell;

Office for Health Improvement and Disparities: John Newton, IQVIA: Helena Jordan, Tim Sheppard, Graham Athey, Dan Moody, Leigh Curry, Pamela Brereton;

National Biocentre: Ian Jarvis, Anna Godsmark, George Morris, Bobby Mallick, Phil Eeles; Glasgow Lighthouse Laboratory: Jodie Hay, Harper VanSteenhouse;

Berkshire and Surrey Pathology Services: Muhammad Ehsaan; Eric Haduli, Hugh Boothe, Reggie Samuel;

Welsh Government: Sean White, Tim Evans, Lisa Bloemberg; Scottish Government: Katie Allison, Anouska Pandya, Sophie Davis;

Public Health Scotland: David I Conway, Margaret MacLeod, Chris Cunningham.

