## Supplement for "Nanopore sequencing of influenza A and B in Oxfordshire and the United Kingdom, 2022-23"

### Nanopore sequencing of influenza A and B viruses in Oxfordshire over the 2022-23 influenza season: supplementary material

#### Supplementary methods

##### Nucleic acid preparation

Nucleic acid was extracted for all samples using the Kingfisher Flex (ThermoFisher) MagMax TaqPath protocol for total nucleic acid isolation from 200µl of sample and eluting in 50µl of elution buffer. Human genomic DNA was removed by DNase treatment with TURBO Dnase (ThermoFisher) at 37°C for 30 minutes. Remaining RNA was purified with RNAClean XP beads (Beckman Coulter) and concentrated into 20µl.

For OUH samples, Ct values obtained by the diagnostic laboratory were used in analyses. For the CIS samples, influenza A and B were differentiated using the RealStar Influenza RT-PCR Kit (Altona Diagnostics) for further processing (see below) and Ct values recorded.

##### Multiplex RT-PCR and purification

For CIS samples, initial PCR testing for influenza A/B was undertaken as follows. Multiplex PCR was performed using Thermo Fisher TaqPath™ COVID-19, Flu A/B, RSV Combo Kit Cat Number A49867. The kit detects multiple viruses: SARS-CoV-2 (N/S genes combined), Influenza A/B (A/B combined), and Respiratory Syncytial Virus (RSV) (A/B combined).
A PCR reaction mixture of 6 µl containing Mastermix and assay was added to a 384-well PCR plate using a multichannel pipette followed by transferring 14 µl of the extracted RNA using the Agilent Bravo Platform. A PCR positive control was added followed by sealing, vortexing, and centrifuging the PCR plate for 2 minutes at 1600 rpm. The PCR plate was run in the Applied Biosystems™ QuantStudio™ 5 Real-Time PCR Instrument according to manufacturer's instructions. The results were analysed using QuantStudio™ Design and Analysis Software version 2.5 and samples were assigned according to the Ct values for the specific target sites as COVID-19, Flu A/B, RSV or combinations of these. Samples positive for influenza A/B were sent to the University of Oxford for targeted RT-PCR for influenza A or B and sequencing. On arrival in Oxford, for the CIS samples, influenza A and B were differentiated using the RealStar Influenza RT-PCR Kit (Altona Diagnostics).

For all samples, purified RNA underwent targeted RT-PCR for influenza A or B. For influenza A, 1µl of each primer were added as detailed previously[1], together with 1µl SuperScript III Platinum Taq (ThermoFisher), 12.5µl 2X Reaction Mix, 3.5µl water and 5µl template RNA. For influenza B, a primer cocktail was made up with previous described primers,[2] with 3µl template RNA. Resulting cDNA was cleaned in a 1:1.8 ratio by Ampure XP (Beckman) bead clean, eluted into 15µl water and quantified by Qubit Fluorometer with the high-sensitivity double-stranded DNA (dsDNA) kit (ThermoFisher).

##### Nanopore library preparation and sequencing

Sequencing libraries were prepared from a total of 4800ng cDNA, multiplexing up to 40 samples per flow cell using Rapid Barcoding 96 Kit SQK-RBK-110.96 (Oxford Nanopore Technologies). Equal cDNA input from all samples was used where possible, however samples with lower cDNA concentrations were included even if below the target cDNA input, using the total input volume for the barcoding reaction, regardless of the quantity of DNA this represented. Libraries were sequenced on FLO-MIN-106 flow cells on a GridION device (Oxford Nanopore Technologies) for 12 hours. Water controls were included for both extraction and RT-PCR steps to confirm no cross contamination. Presence of cDNA fragments in these control samples was also assessed by TapeStation platform (Agilent).

##### Illumina sequencing

Illumina sequencing was undertaken to evaluate the accuracy of Nanopore sequencing. Purified RNA from influenza A positive samples underwent RT-PCR as previously described.[3] Resulting cDNA was quantified as above. Nextera XT DNA Library Prep kit (Illumina) was used following manufacturer's instructions through tagmentation and a 12 cycle indexing PCR. The resulting cDNA was cleaned with Ampure XP beads in a 1:0.6 ratio and then quantified by Qubit. Libraries were normalised to 4nM, pooled, denatured with 0.2N NaOH and PhiX was added at 1% total volume. Sequencing was run for 300 cycles using a MiSeq Reagent Kit V2.

##### Bioinformatic and statistical analysis

The bioinformatics workflow was written in nextflow[4] and packaged in conda, it is publicly available on gitlab here https://gitlab.com/ModernisingMedicalMicrobiology/flu-workflow. Nanopore sequence reads were basecalled and demultiplexed using Guppy version 6.5.7+ca6d6af with default settings for the library preparation used. Sequenced reads were simultaneously competitively mapped using minimap2 v2.24-r1122 (with -ax ont settings) to reference genomes representing each of the major influenza A and B subtypes: A/California/07/2009(H1N1), A/Shanghai/02/2013(H7N9), A/Korea/426/1968(H2N2), A/Puerto Rico/8/1934(H1N1), A/New York/392/2004(H3N2), A/Goose/Guangdong/1/96(H5N1), A/Hong Kong/1073/99(H9N2), B/Lee/1940, B/Victoria/02/1987, B/Yamagata/16/1988. Accessions for reference genomes are available in the workflow gitlab repository. Aligned bam files were generated from filtered minimap2 output using samtools v1.16.1 (with -F 2304 and -q 50 settings). Variant calling was performed with clair3[5] and applied to generate consensus sequences with bcftools (v1.18) with a minimum depth of 10x reads.

Each reference genome contains 8 segments. To address potential misidentification of samples arising from low level laboratory contamination, barcode cross-over effects arising from imperfect demultiplexing of reads, and Nanopore sequencing errors, we developed several filters, as previously developed approaches, e.g., requiring 20-fold coverage (https://github.com/epi2me-labs/wf-flu) of each segment were not optimally sensitive or specific.

Our filters were used for both influenza A and B to confirm the presence within a sequenced sample of a specific segment of a given reference genome, e.g., A/H3N2 segment 4 (S4, representing the haemagglutinin gene [HA]). We used influenza A sequences to derive these filters. We assumed that mixed infections were likely to be uncommon (with the exception of recent live attenuated influenza vaccine [LAIV] recipients). For each sample and reference genome segment without evidence of recent LAIV exposure, we identified the subtype with the greatest number of mapped reads, e.g., for HA, H3. We then considered all reads from this sample that mapped to the same segment, HA, but to other subtypes, e.g., H1, H2, etc.

Assuming these represented misclassified reads, we visualised the total number of misclassified reads within the sample and the percentage these reads represented out of all reads across all samples in a sequencing run (i.e., same flow cell) mapped to that segment. The latter filter was introduced to account for barcode misclassification leading to apparent contamination where another sample in the sequencing run of a different subtype had very high coverage. Using sequencing runs conducted up to 12 April 2023, by inspecting these distributions for the HA and NA segments (segments 4 and 6) (Figure S1 and Figure S2), we set requirements for ≥10 reads to be mapped to a segment and ≥0.5% of all reads in a run to be mapped to a segment (typically up to 30 samples were multiplexed per run, so the expected value with even coverage would be approximately 3%). Additionally, to address low level cross subtype mapping of reads arising from similarities between the same segment in different subtypes and Nanopore sequencing errors, we required ≥1% of all reads mapped to any subtype of a segment within a sample to be mapped to each identified subtype. As such we could only identify mixed infections where the minority subtype accounted for ≥1% of the sequenced reads. Finally, we also required ≥60% of the length of the identified segment to have ≥10 fold depth. By design, our approach means that low level mixed infections are excluded, but genuine mixed infections at even modest levels could still be identified. Our approach is also robust to changes in the depth of sequencing, whereas a single absolute depth threshold fails at very high sequencing depth, due to the possibility of misclassification described above.

Having developed these filters on segments 4 and 6, we then applied these filters to all other segments. For segments 2 and 3 (S2 and S3), cross subtype mapping was much more common and so these segments were not used to identify viral subtypes or mixed infections. Probable LAIV samples were identified by the presence of H2N2 segments 1, 5, 7 and 8, together with segments 4 and 6 from both H1N1 and H3N2. For all other samples, we used the presence of at least 2 segments excluding S2 and S3 of the same influenza subtype to confirm identification of a subtype. Hence even if segments 4 and 6 (HA and NA genes) were not successfully sequenced we could still infer e.g. A/H3N2 as being present based on other segments in circulating subtypes.

Phylogenetic trees were created based on alignments of all successfully sequenced HA segments for A/H3N2, A/H1N1, and B/Victoria sequences. As consensus input sequences could include insertions and deletions, alignments were produced using mafft (v7.508 with –auto settings) and maximum likelihood trees generated using raxmlHPC-PTHREADS-AVX2 (version 8.2.12; with -f a -m GTRGAMMA -p 12345 -x 12345 -# 10 settings). Strains for 2022-23 northern hemisphere vaccines[6] were included in the trees, with one randomly selected as an outgroup to root the tree. Similarly, phylogenetic trees were created based on NA segments, and for whole genomes. Where segments were not successfully sequenced, these were included in whole genome trees as Ns, with an additional requirement for ≥60% of the whole genome to be called for a sequence to be included. As we noted that segments S2 and S3 were more prone to cross-sample mapping (see above), as a sensitivity analysis we also created trees based on the six remaining segments. Pairwise SNPs between segment and whole genome sequences were obtained from the cophenetic distances extracted from trees multiplied by the alignment length, and as such account for variable amounts of missingness in the proportion of each segment or genome called.

Rates of influenza HA segment mutation were estimated from sample times and sequence alignments using BEAST (version 1.10.4). An HKY substitution model and strict molecular clock were assumed. Default priors were used. Four chains were run for each of A/H3N2 and A/H1N1 and checked to ensure convergence. Each chain was run for ≥10 million iterations, after discarding ≥10% of the iterations as burn-in, analyses were run for sufficient iterations to ensure an effective sample size of >200 for all parameters. There were insufficient samples to undertake this analysis for B/Victoria.

To minimise laboratory contamination, any sequencing run with any segment of any influenza type identified in a water control was discarded. To ensure reproducibility of sequencing and sample handling, at least 1 previously sequenced sample was included on each sequencing run. If any of the replicate sequences were assigned mis-matching influenza types, e.g. A/H3N2 on one run and A/H1N1 on another, or if the number of single nucleotide polymorphisms (SNPs) between replicate HA segment sequences excluded a plausible maximum of 10 SNPs (see results), then both runs were discarded and repeated where sufficient sample volume remained.

IRMA (https://wonder.cdc.gov/amd/flu/irma/) was used to assemble raw Illumina reads into consensus sequences for each segment. Default options were used and only the coding regions of each segment were assembled. Only segments with greater than 60% of the bases called were kept for analysis. The A/H3N2 segment 4 was extracted from each nanopore or Illumina assembly and aligned using biopython[7] ( pairwise2.align.globalms(seq1, seq2, 2, -1, -3, -2) ) with a higher gap open penalty to preserve SNP positions relative to each segment. Aligned positions were iterated and compare if the base present was not uncalled ('N') or missing and represented with a gap ('-').

#### Supplementary figures

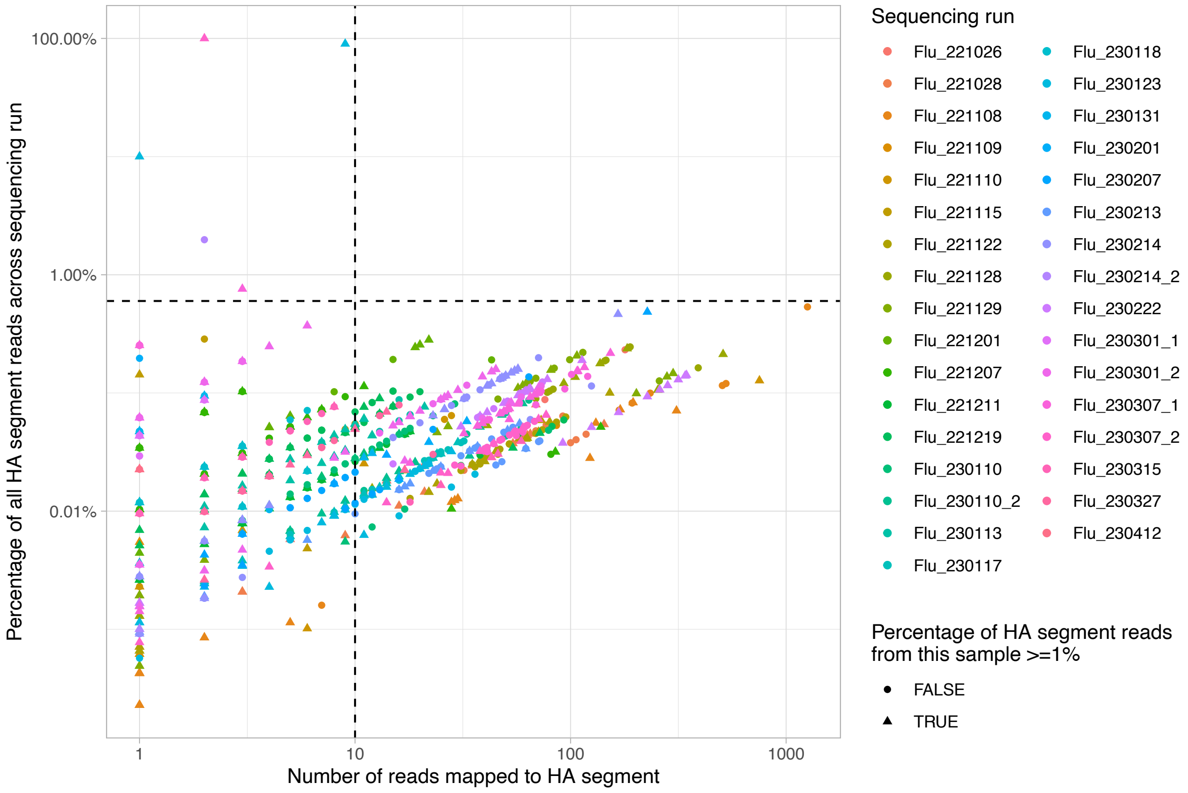


**Figure S1. Influenza segment 4 (HA gene) misclassification**. The dashed thresholds and shapes indicate those chosen for excluding misclassification.

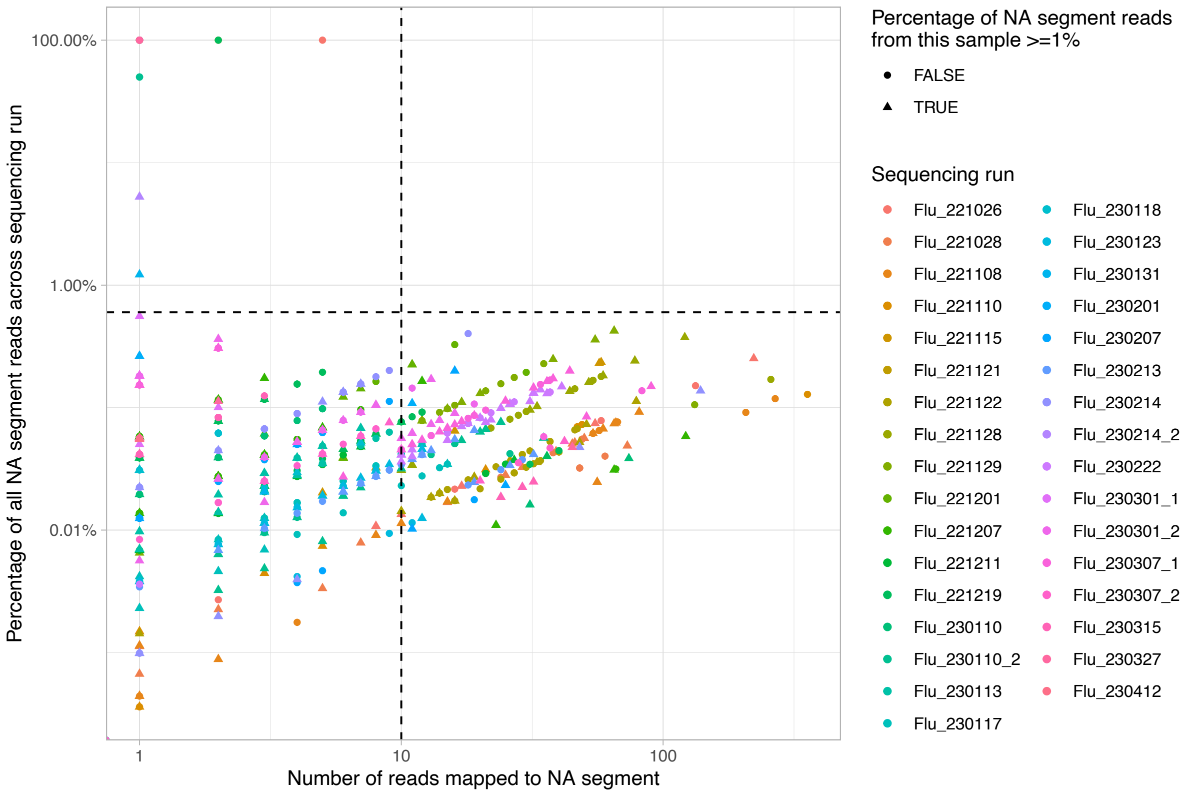


**Figure S2. Influenza segment 6 (NA gene) misclassification**. The dashed thresholds and shapes indicate those chosen for excluding misclassification.

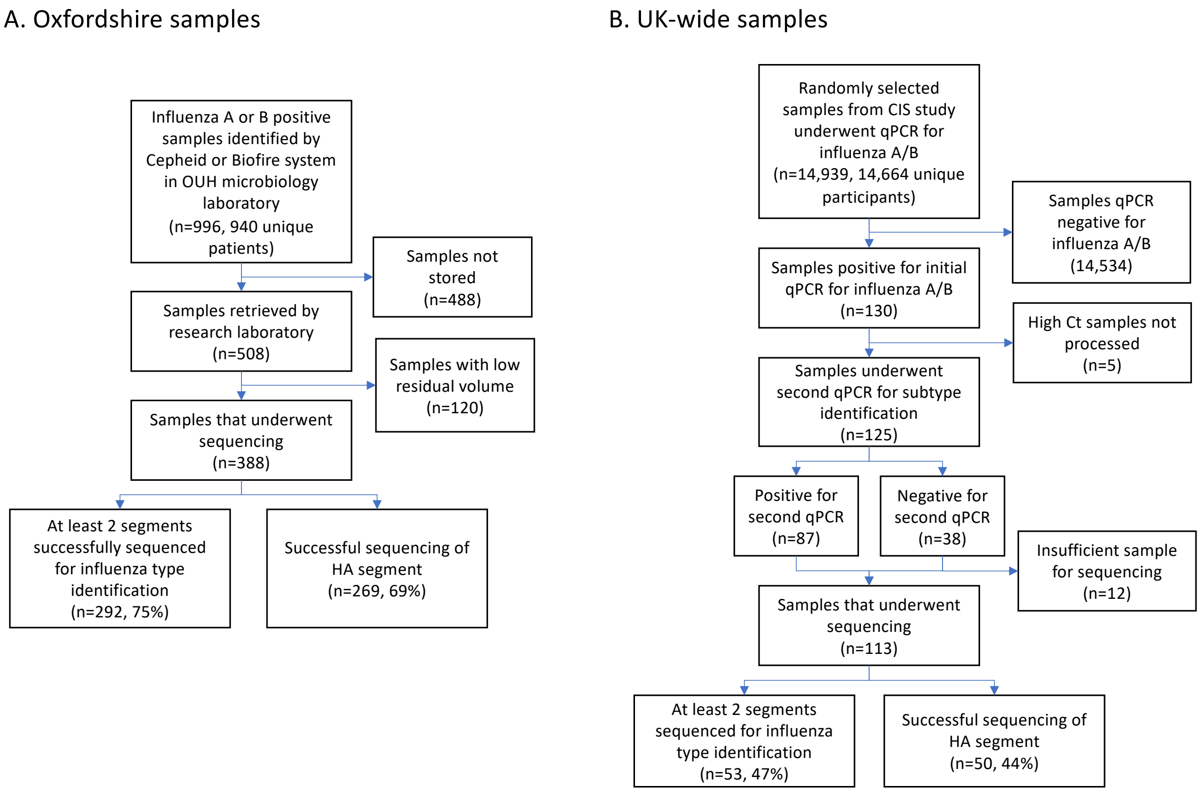


**Figure S3. Samples collected and sequenced**. OUH, Oxford University Hospitals. CIS, Office for National Statistics COVID-19 Infection Survey. Ct, cycle threshold.

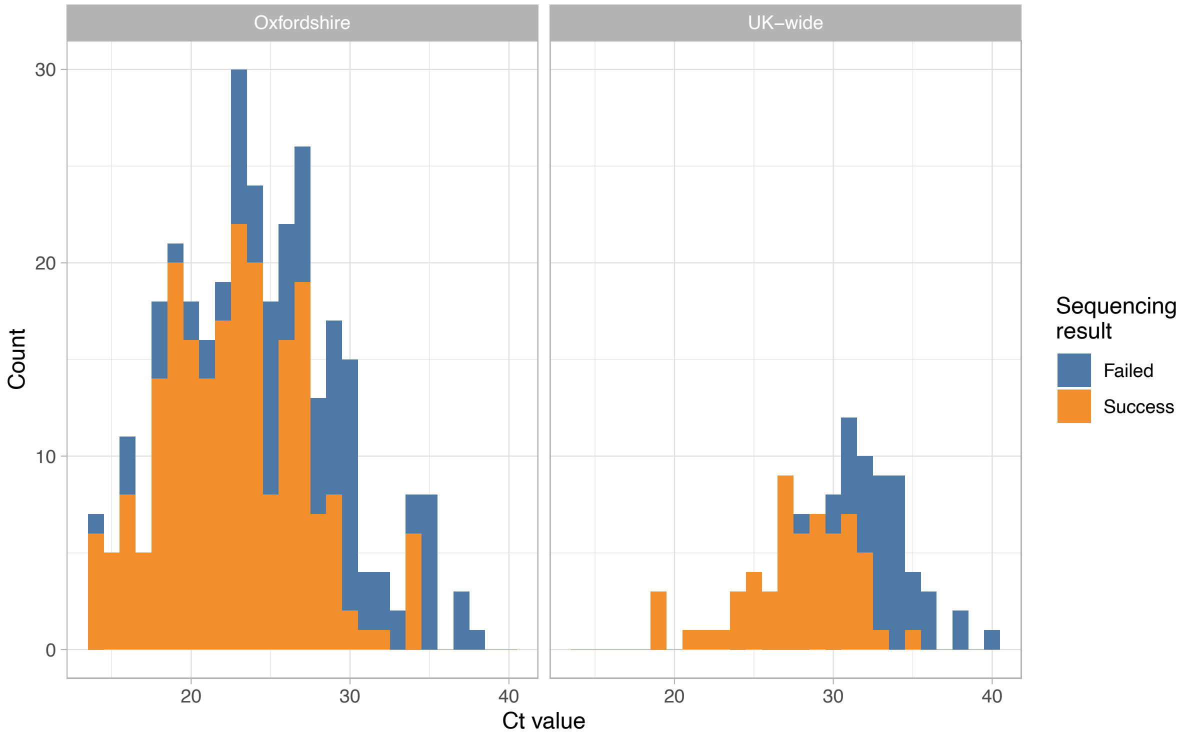


**Figure S4. HA segment sequencing success by Ct values and study.**

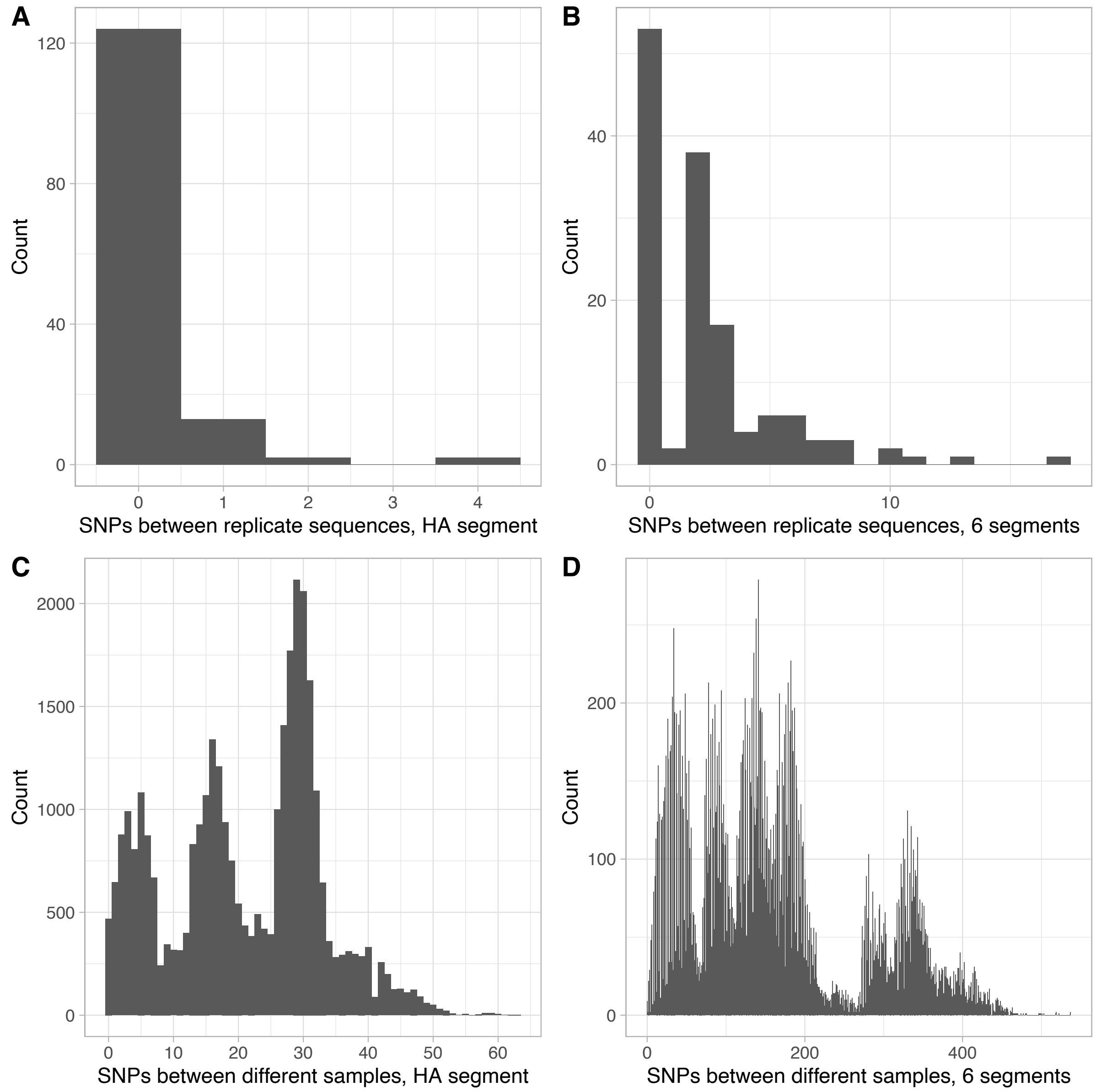


**Figure S5. Distribution of single nucleotide polymorphism (SNPs) between replicate and different sample pairs**. Panels A (n=141) and C (n=32,992) show differences for the HA segment and B (n=137) and D (n=29,398) for genomes based on six segments (segments S2 and S3 were omitted because of a greater tendency to cross mapping between flu types, see Figure 2 for whole genome comparison).

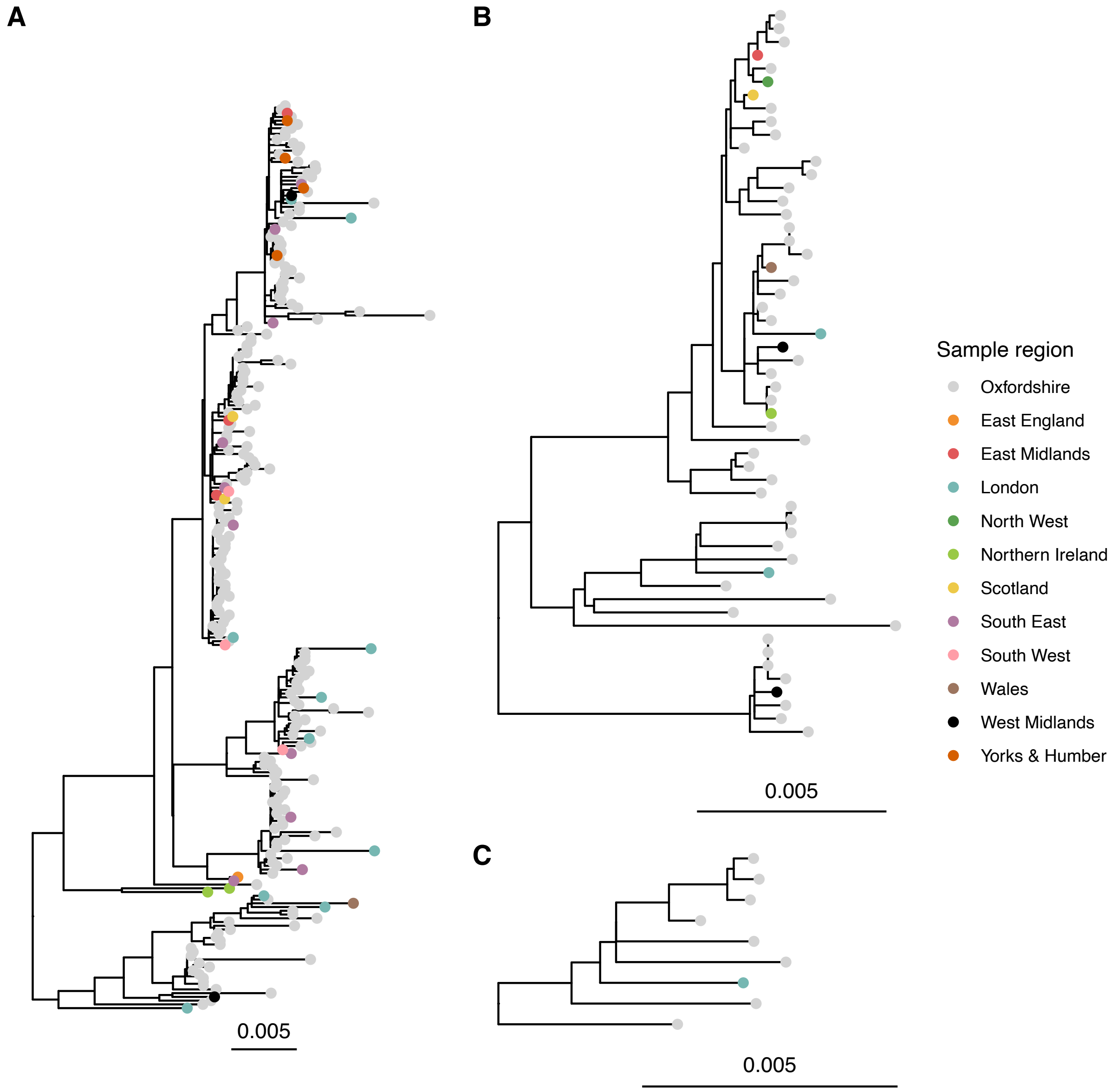


**Figure S6. Unrooted maximum likelihood phylogeny of six segment sequences for influenza A H3N2 (panel A), H1N1 (panel B), and influenza B/Victoria (panel C).** Segments S2 and S3 were excluded from whole genome alignments due to a higher propensity to be impacted by cross sample mapping.
